# Uveoscleral Outflow Enhancement as a Complementary Bio-intervention after Failed Prior Trabecular Outflow Treatment: 12-Month Outcomes of Allograft-reinforced Cyclodialysis in Open Angle Glaucoma

**DOI:** 10.64898/2026.11.12.26367933

**Authors:** Gus Gazzard, Hari Jayaram, Reena Garg, Tsontcho Ianchulev, Robert Stamper, James Tsai, I. Paul Singh

**Affiliations:** Moorfields Eye Hospital, 162 City Road, London, EC1V 2PD, UK; Georgetown University Hospital, 3800 Reservoir Rd NW, Washington, DC, 20007, USA; New York Eye and Ear Infirmary of Mount Sinai, 310 E 14th Street, New York, NY, 10003, USA; University of California, San Francisco, 490 Illinois Street, San Francisco, CA, 94158, USA; Eye Centers of Racine and Kenosha, 13200 Globe Drive, Suite 216, Mt. Pleasant, WI, 53177, USA

**Keywords:** glaucoma, open-angle, cyclodialysis clefts, intraocular pressure, minimally invasive surgical procedures, ophthalmologic surgical procedures

## Abstract

**Background:** Eyes with open-angle glaucoma uncontrolled after a trabecular outflow procedure have limited options before progressing to bleb-forming surgery. The uveoscleral pathway is distinct, relatively pressure-independent, and conjunctiva-sparing. We report 12-month outcomes of standalone allograft-reinforced cyclodialysis in eyes with IOP that remained uncontrolled after prior trabecular outflow intervention.

**Methods:** Analysis of a multicentre registry (CREST; NCT05506423). Eligible eyes had at least one prior trabecular outflow procedure, no prior bleb-forming surgery, and underwent uveoscleral outflow bio-intervention with at least 12 months of follow-up. IOP reported as mean with 95% CI, medication as mean ± SD.

**Results:** Forty-seven eyes were eligible; 91.5% completed 12 months. Consistent with moderate-to-advanced glaucoma, baseline IOP was 22.5 mmHg (20.1, 24.9) on 3.0 (1.3) IOP-lowering medications, with visual field mean deviation −11.7 dB (8.1). 95.7% were pseudophakic. In the per-protocol efficacy analysis, IOP fell 6.1 mmHg (3.7, 8.6), a 25.0% reduction (15.5%, 34.5%; p < 0.001), with no increase in medication. At 12 months, the Kaplan–Meier estimate of freedom from incisional reoperation was 78.2% (61.8%, 88.2%). There were no vision-threatening, clinically significant adverse events associated with the procedure such as persistent hypotony, maculopathy or other complications.

**Conclusions:** In eyes with inadequately controlled IOP following prior trabecular intervention, standalone allograft-reinforced cyclodialysis was associated with clinically meaningful IOP reduction without an increase in medication burden or permanent vision loss.

## 1. Introduction

Lowering intraocular pressure (IOP) remains the only intervention proven to slow glaucomatous progression^1^. The conventional (trabecular) outflow pathway, comprising the trabecular meshwork, Schlemm canal, collector channels and the episcleral venous system, carries the majority of aqueous and hosts the principal sites of pathological resistance in open-angle glaucoma (OAG)^2,3^. The unconventional (uveoscleral) pathway drains across the ciliary muscle face into the supraciliary and suprachoroidal spaces and is comparatively pressure-independent, functioning as a relief route whose capacity rises as trabecular resistance increases^3^. Pharmacologically, this pathway is the target of the most effective class of ocular hypotensive medication: prostaglandin analogues lower IOP principally by enhancing uveoscleral outflow^4^ and produce the largest IOP reduction of any first-line medication^5,6^.

This distinction has a direct therapeutic consequence. Trabecular-based minimally invasive glaucoma surgery (MIGS) can only reduce IOP down to the pressure set by distal resistance and episcleral venous pressure, which in practice imposes a post-trabecular IOP floor in the mid-teens^7,8^. A level-I American Academy of Ophthalmology assessment of randomised data found that adding a trabecular procedure to cataract surgery yields only 1.6 to 2.3 mmHg (3.8% to 8.9%) of additional IOP reduction beyond the 5.4 to 7.6 mmHg (21% to 28%) provided by phacoemulsification itself^9^. Approximately three-quarters of the IOP reduction seen in combined cataract-MIGS is therefore attributable to the cataract extraction rather than the trabecular device. When the lens contribution is removed, as in the pseudophakic, standalone setting, the ceiling of trabecular surgery becomes clinically apparent, and once that pathway has already been surgically manipulated and has failed, further trabecular intervention offers diminishing physiological returns.

For a pseudophakic eye with moderate-to-advanced glaucoma that remains uncontrolled after a prior trabecular procedure, this leaves a therapeutic gap. The available options have historically been either another attempt at the same, already-compromised conventional pathway or an escalation to bleb-forming surgery. Bleb-forming surgery is the most effective IOP-lowering approach but carries a lifelong risk of bleb-related complications, such as infection, hypotony and scarring-related failure, and consumes conjunctival tissue whose prior disturbance measurably lowers the success of any subsequent filtration procedure^1,10,11^. A conjunctiva-sparing option that addresses the untouched, complementary outflow pathway is therefore a useful intermediate step.

The uveoscleral (unconventional) outflow pathway is a physiologically distinct target. It accounts for 35%-50% of total aqueous outflow in healthy adults and functions as a relatively pressure-independent route where aqueous humour traverses the ciliary muscle into the supraciliary and suprachoroidal spaces^12,13^. Prostaglandin analogues act predominantly along this pathway, lowering IOP through remodelling of ciliary muscle extracellular matrix^4,14^. Surgical attempts to harness the uveoscleral pathway date back to Heine’s cyclodialysis procedure in 1905. Despite its potent IOP-lowering potential, classic cyclodialysis fell out of favour due to unpredictable cleft closure and hypotony. A recent systematic review and meta-analysis of over 4,000 glaucoma cases spanning 100 years of surgical experience found cyclodialysis to be effective in lowering IOP as a uveoscleral outflow-enhancing procedure, with modern ab-interno approaches showing improved efficacy, safety and procedural longevity compared with historical ab-externo techniques^15^. Early attempts to stent the supraciliary space with synthetic materials (e.g., the CyPass Micro-Stent) confirmed the hydraulic efficacy of the pathway as a potent outflow channel, similar to the high therapeutic index seen with uveo-scleral IOP lowering agents such as prostaglandin analogues^16,17^.

Allograft-reinforced cyclodialysis is a newer approach to uveoscleral outflow enhancement. An allogeneic, acellular scleral bio-scaffold reinforces the cyclodialysis cleft, with the aim of promoting bio-integration instead of the fibrous encapsulation seen with synthetic supraciliary devices^18,19^. This report presents 12-month outcomes of allograft-reinforced cyclodialysis performed as a standalone procedure in eyes with a history of prior trabecular intervention and inadequate IOP control on medical therapy, in a predominantly pseudophakic population.

## 2. Methods

### Study Design and Population

The CREST Study (NCT05506423) is a multicentre, observational, real-world evidence registry, in which patients with OAG who underwent bio-interventional uveoscleral enhancement surgery with the AlloFlo Uveo™ System (Iantrek, Inc., White Plains, NY) are prospectively followed through 24 months postoperatively. CREST is conducted in accordance with the Declaration of Helsinki, with approval from relevant Institutional Review Boards for all participating investigators. All participants provided written informed consent prior to enrolment. This study analysed a subgroup of eyes from CREST Cohort 2 (allograft-reinforced cyclodialysis) meeting the following inclusion criteria: (1) history of at least one prior trabecular outflow intervention (defined as any of canaloplasty, trabecular stent implantation, goniotomy, trabeculotomy, or laser trabeculoplasty), (2) no prior conjunctival bleb-forming procedure (trabeculectomy, tube shunt, subconjunctival gel stent), (3) placement of two allogeneic scleral allografts during a single surgical session, and (4) the surgery had been performed at least 12 months before data cutoff for analysis. As an uncontrolled subgroup analysis of a single-arm registry, this study is not designed to support comparative claims against alternative interventions.

### Surgical Procedure

Two minimally modified, homologous allograft spacers of processed, acellular scleral tissue (5 mm × 500 µm) were deployed into the supraciliary space to provide structural support and serve as a porous conduit for aqueous humour drainage into the uveoscleral pathway. The allograft-reinforced cyclodialysis procedure was performed using an ab-interno approach under gonioscopic visualisation. Elongated acellular scleral allograft tissue constructs were prepared and introduced into the anterior chamber (Figure 1) under gonio-visualisation (Figure 2). The allografts were deployed at the endoscleral surface bilaterally and adjacent to the edges of the cyclodialysis. Components did not protrude into the anterior chamber (AC), and AC-clear placement was confirmed under direct visualisation with the allografts deployed flush with the iris root. Any residual viscoelastic was evacuated with irrigation or irrigation/aspiration. Patients were prescribed a surgeon’s choice of postoperative antibiotics and corticosteroids with taper.

**Figure 1.**
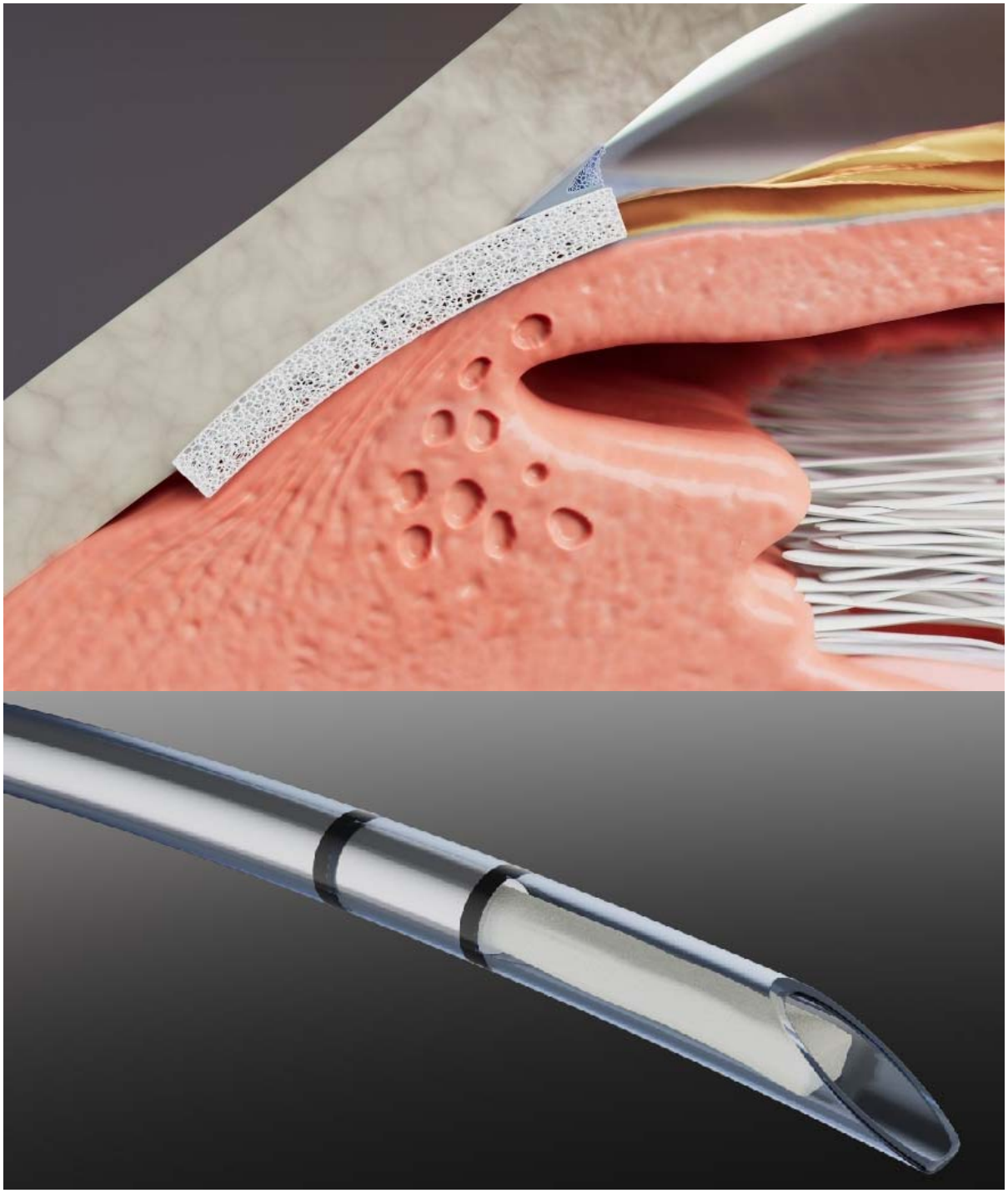
Placement of Allograft Reinforcement. (Top) Bio-interventional cyclodialysis with the allograft tissue deployed for reinforcement. (Bottom) Sleeved interventional deployment with a cyclodialysis cannula.

**Figure 2.**
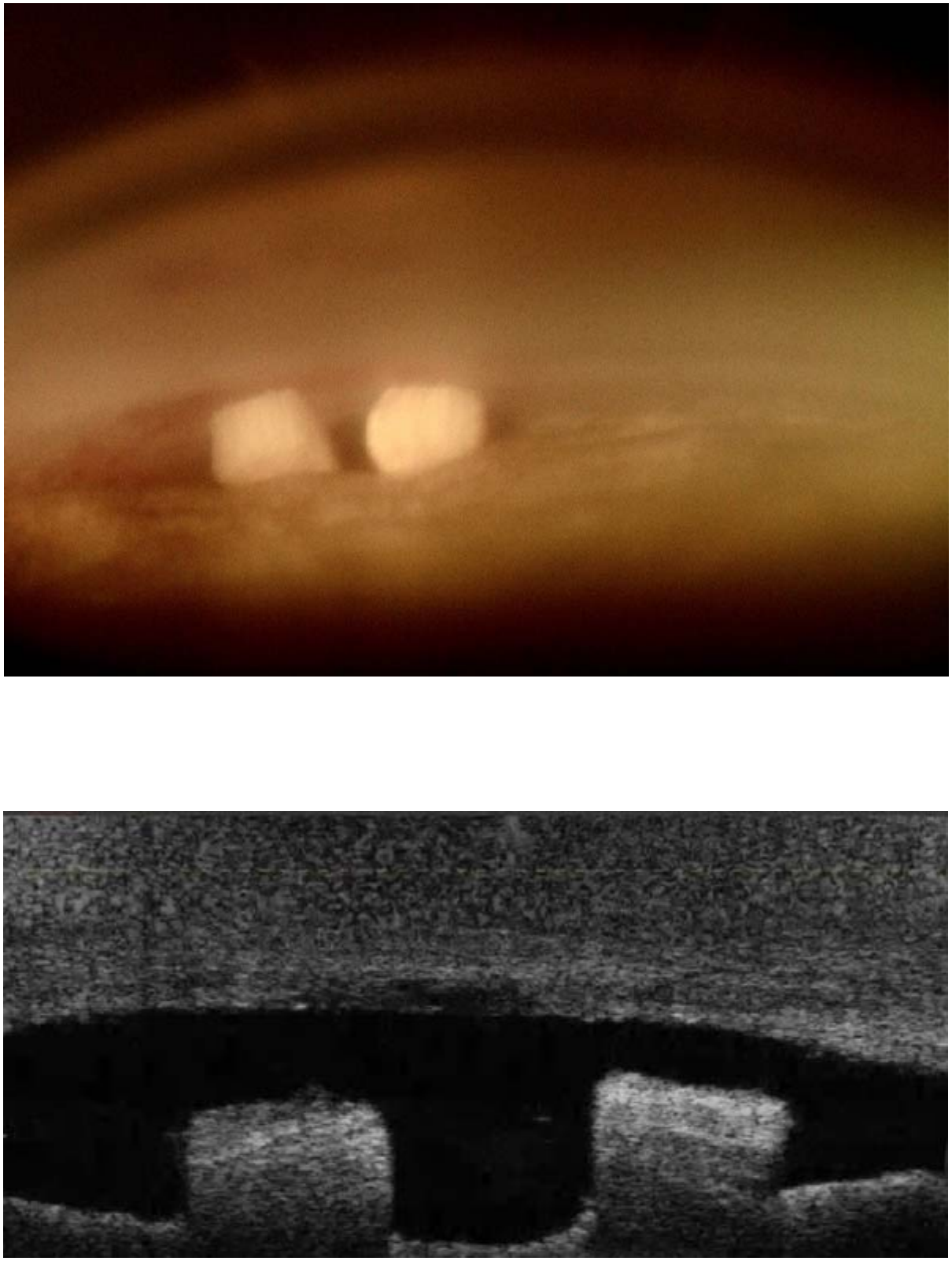
In-Situ Images of Allogeneic Tissue. Two acellular scleral allografts in situ at ≥ 90 days after standalone bio-reinforced cyclodialysis. (Upper) Scleral transillumination, with the globe backlit so that the allografts appear as discrete light-transmitting blocks within the cleft. (Lower) Anterior-segment optical coherence tomography (OCT) through the same region, showing both allografts spanning the cyclodialysis cleft with a patent, optically clear space maintained between the sclera and the ciliary body.

### Outcome Measures

Efficacy data were collected at baseline and at postoperative Months 1, 6, and 12. Key efficacy endpoints of interest included: mean medicated IOP over time, mean change in IOP from baseline to Month 12, mean change in medication burden, and responder rates. Safety endpoints included the incidence of ocular adverse events, specifically clinically significant hypotony (prespecified as IOP < 6 mmHg with clinical sequelae), IOP elevations ≥ 10 mmHg from baseline occurring > 30 days postoperatively, postoperative hyphaema > 2 mm, allograft migration or extrusion, ciliary or choroidal effusion, and reoperation.

### Statistical Analysis

Longitudinal changes in IOP and medication burden from baseline were tested using a paired t-test. Statistical significance was defined as p < 0.05. IOP data are presented as means with 95% confidence intervals (95% CI); medications as means ± standard deviation (SD). Responder rates are reported at IOP thresholds of ≤ 21, ≤ 18 and ≤ 15 mmHg and at ≥ 20% reduction from baseline. Composite responder was defined as meeting the stated IOP threshold without an increase in glaucoma medications from baseline. Time to reoperation was estimated by Kaplan-Meier analysis with complementary log-log confidence intervals. Reoperation was defined as any incisional IOP-lowering procedure, and eyes without a reoperation were censored at their last scheduled study visit. In a sensitivity analysis, any subsequent IOP-lowering procedure, including cyclophotocoagulation, laser trabeculoplasty and laser peripheral iridotomy, was counted as a failure; anterior chamber paracentesis was not.

For each protocol visit, mean IOP and mean medication burden were computed using all eyes with a measurement available in that visit’s window, with per-visit N reported explicitly; no imputation was applied. Consistent with reporting conventions for real-world registry analyses of glaucoma surgery outcomes^20^, eyes were censored from IOP and medication summary analyses at the time of any subsequent IOP-lowering procedure. Analyses were performed using R version 4.5.3 (R Project for Statistical Computing).

## 3. Results

### Demographics and Baseline Disease Characteristics

A total of 47 eyes (45 patients) met all inclusion criteria and were included for analysis (Table 1). Thirty-two eyes had a Month 12 pressure recorded before undergoing any further pressure-lowering procedure. The remainder had already undergone a further procedure, so their later readings reflect that intervention rather than the index surgery.

**Table 1.** Summary of Demographics and Baseline Disease Characteristics.

| **Patient Demographics** | **Patients (N=45)** |
|---|---|
| Age, mean years (SD) | 74.2 (10.2) |
| Female, n (%) | 24 (53.3%) |
| Non-white, n (%) | 15 (33.3%) |
| Hispanic or Latino, n (%) | 6 (13.3%) |
| **Baseline Disease Characteristics** | **Eyes (N=47)** |
| Medicated IOP, mean mmHg (95% CI)□ | 22.5 (20.1, 24.9) |
| IOP-lowering medications, mean n (SD) | 3.0 (1.3) |
| 0 | 2 (4.3%) |
| 1 | 3 (6.4%) |
| 2 | 13 (27.7%) |
| ≥ 3 | 29 (61.7%) |
| Visual field mean deviation, mean dB (SD)□ | −11.7 (8.1) |
| Lens status, n (%) |  |
| Pseudophakic | 45 (95.7%) |
| Phakic | 1 (2.1%) |
| Aphakic | 1 (2.1%) |
| Baseline gonioscopy grade, n (%) |  |
| Grade 4 | 20 (42.6%) |
| Grade 3 | 4 (8.5%) |
| Grade 2 | 1 (2.1%) |
| Not recorded | 22 (46.8%) |
| Systemic anticoagulant/antiplatelet, n (%) | 5 (10.6%) |
| Prior Glaucoma Procedures, n (%)□ |  |
| Number of prior glaucoma procedures |  |
| 1 | 16 (34.0%) |
| 2 | 18 (38.3%) |
| 3 | 7 (14.9%) |
| ≥ 4 | 6 (12.8%) |
| Prior interventions performed |  |
| Canaloplasty | 24 (51.1%) |
| Laser trabeculoplasty | 37 (78.7%) |
| Trabecular stent implant | 9 (19.1%) |
| Goniotomy/trabeculotomy | 7 (14.9%) |
| Bleb-forming procedure | 0 (0%) |

CI = confidence interval; IOP = intraocular pressure; SD = standard deviation.
a. Baseline IOP available for all 47 eyes.
b. Baseline visual field mean deviation available for 38 eyes.
c. Eyes could have had more than one prior procedure, so categories are not mutually exclusive. Bleb-forming surgery was an exclusion criterion.

The cohort was an advanced glaucoma population that had failed at least one prior canal/trabecular intervention: mean baseline medicated IOP was 22.5 mmHg (20.1, 24.9); eyes were on a mean (SD) 3.0 (1.3) topical IOP-lowering medications, with 61.7% on ≥ 3 medications; mean (SD) visual field mean deviation was −11.7 (8.1) dB. Most eyes were pseudophakic (45 [95.7%]); one eye was phakic and one aphakic. Five eyes (10.6%) were taking a systemic anticoagulant or antiplatelet agent.

At baseline, eyes had undergone the following prior trabecular interventions: laser trabeculoplasty (78.7%), canaloplasty (51.1%), trabecular stent implantation (19.1%), and goniotomy/trabeculotomy (14.9%). Sixty-six per cent had undergone two or more prior trabecular or angle-based procedures and 27.7% three or more, counting each component of a combined procedure separately. Baseline gonioscopy grade, where recorded, was Grade 4 in 20 eyes (42.6%), Grade 3 in 4 (8.5%) and Grade 2 in 1 (2.1%); grade was not recorded in 22 eyes (46.8%). Peripheral anterior synechiae were not captured as a discrete baseline field in the registry and could not be summarised.

### Efficacy Outcomes

Mean medicated IOP decreased from 22.5 mmHg (20.1, 24.9) at baseline (N=47) to 20.3 mmHg (16.5, 24.2) at Month 1 (N=42), 16.3 mmHg (13.8, 18.8) at Month 6 (N=37), and 14.7 mmHg (12.7, 16.7) at Month 12 (N=32) (Figure 3, Table 2). Among the 32 eyes whose Month 12 pressure preceded any further procedure, mean IOP change from baseline was −6.1 mmHg (−8.6, −3.7), a relative reduction of 25.0% (15.5%, 34.5%; p < 0.001). At Month 12, 96.9% of these eyes had IOP ≤ 21 mmHg, 87.5% ≤ 18 mmHg and 68.8% ≤ 15 mmHg; 68.8% achieved a ≥ 20% reduction and 40.6% a ≥ 30% reduction (Figure 4). Composite responder rates, meeting the IOP threshold with no increase in medication from baseline, were 23 eyes (71.9%) for IOP ≤ 21 mmHg, 22 (68.8%) for ≤ 18 mmHg, 18 (56.2%) for ≤ 15 mmHg (Figure 4, Table 2). In a sensitivity analysis restricted to eyes whose prior intervention included an incisional or angle-based procedure rather than laser alone (35 eyes), the Month 12 reduction was 5.8 mmHg (2.8, 8.7), or 22.5% (11.4%, 33.6%; p < 0.001; paired N=25).

**Figure 3.**
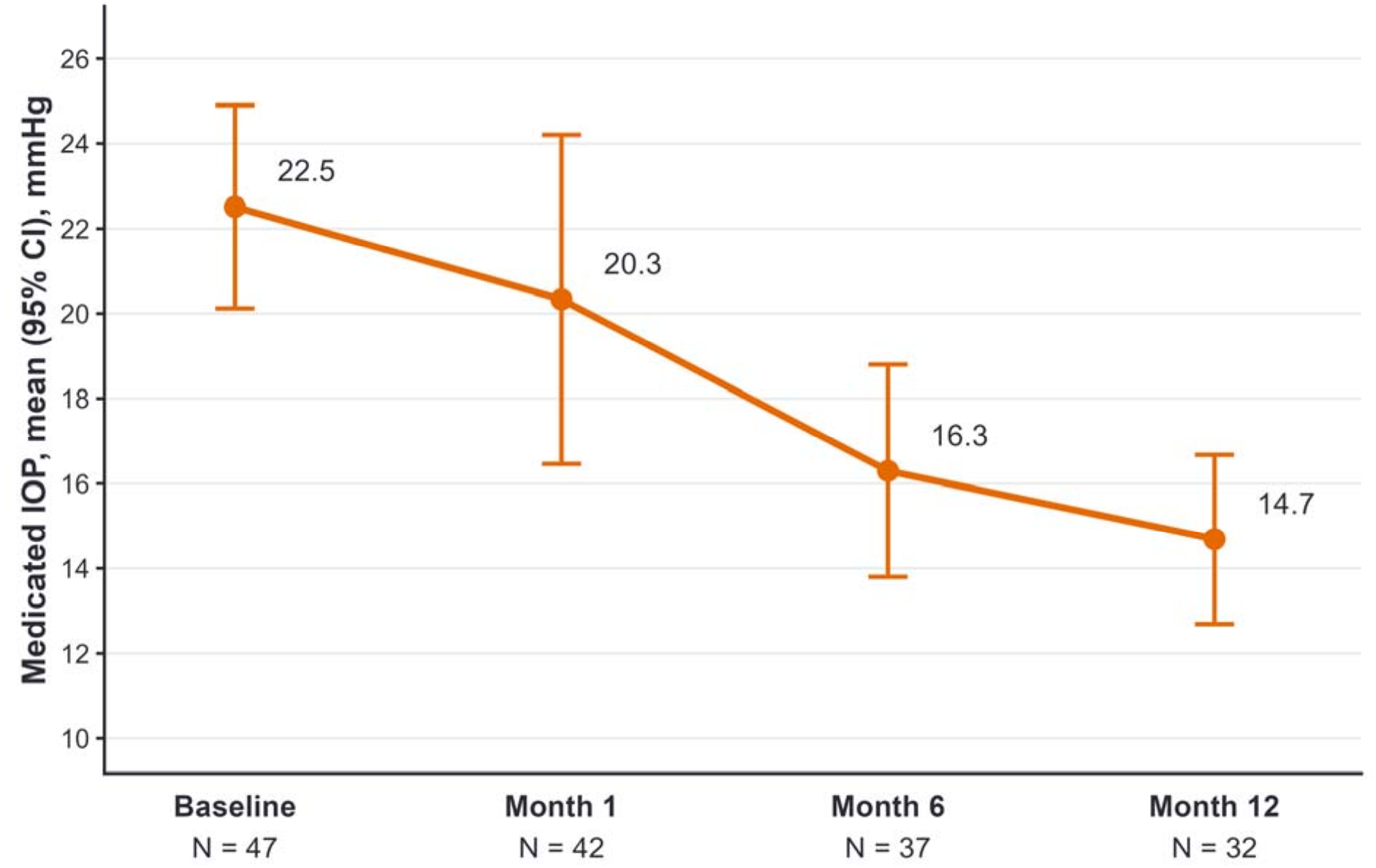
Mean IOP Over Time. CI = confidence interval; IOP = intraocular pressure. Note: values reflect all eyes with an available measurement at each timepoint, with observations censored after any subsequent IOP-lowering procedure: N=47, 42, 37 and 32 at baseline and Months 1, 6 and 12 respectively.

**Figure 4.**
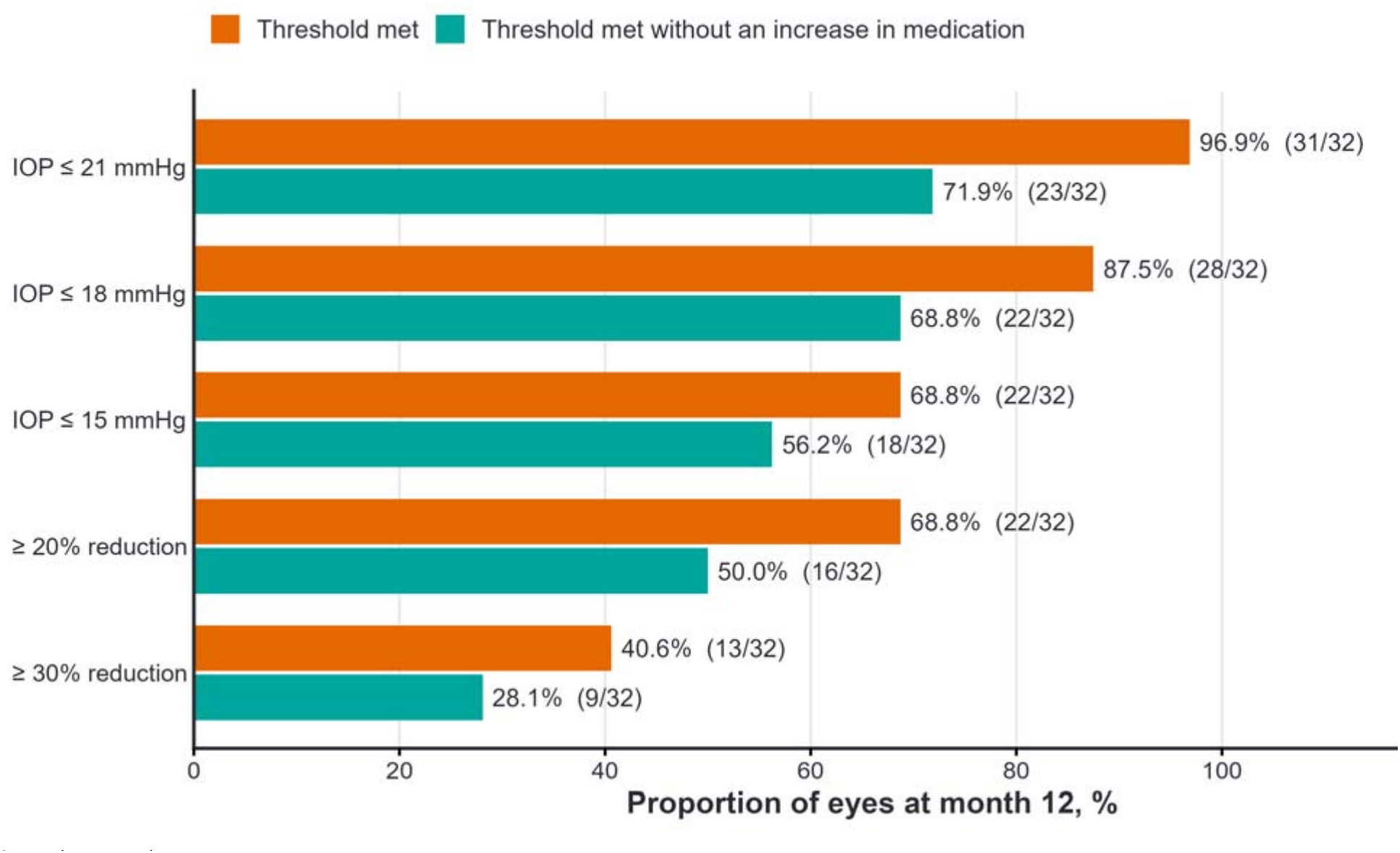
Proportion of Eyes Achieving Intraocular Pressure Thresholds and Responder Criteria at Month 12. IOP = intraocular pressure. Notes: For each criterion, bars show the proportion of eyes meeting it (orange) and the proportion meeting it without an increase in glaucoma medications from baseline (green). Proportions are among the 32 eyes whose Month 12 pressure preceded any further procedure.

**Table 2.** Summary of Efficacy Outcomes.

| Efficacy Outcome | Analysis cohort<br>(N=47) |
|---|---|
| **Mean IOP over time, as observed after censoring, mmHg (95% CI)** |  |
| Baseline (N=47) | 22.5 (20.1, 24.9) |
| Month 1 (N=42) | 20.3 (16.5, 24.2) |
| Month 6 (N=37) | 16.3 (13.8, 18.8) |
| Month 12 (N=32) | 14.7 (12.7, 16.7) |
| **Mean IOP change from baseline to Month 12, with censoring (paired N=32)** |  |
| Baseline, mmHg (95% CI) | 20.8 (18.3, 23.3) |
| Month 12, mmHg (95% CI) | 14.7 (12.7, 16.7) |
| Absolute change, mmHg (95% CI) | −6.1 (−8.6, −3.7) |
| Relative change, % (95% CI) | −25.0% (−34.5%, −15.5%) |
| p-value | p < 0.001 |
| **Mean medication burden, as observed over time, n (SD)** |  |
| Baseline (N=47) | 3.0 (1.3) |
| Month 1 (N=42)* | 2.1 (1.6) |
| Month 6 (N=37) | 2.7 (1.7) |
| Month 12 (N=32) | 2.8 (1.5) |
| **Mean medication change from baseline to Month 12 (paired N=32)** |  |
| Baseline, n (SD) | 3.0 (1.3) |
| Month 12, n (SD) | 2.8 (1.5) |
| Change from baseline, n (95% CI) | −0.1 (−0.50, 0.25) |
| p-value | p = 0.50 |
| **Proportion of medication-free eyes, as observed over time, n (%)** |  |
| Baseline (N=47) | 2 (4.3%) |
| Month 1 (N=42)* | 7 (16.7%)* |
| Month 6 (N=37) | 4 (10.8%) |
| Month 12 (N=32) | 1 (3.1%) |
| **Sensitivity analysis: mean IOP in eyes with a prior incisional or angle-based procedure (paired N=25)** |  |
| Baseline, mmHg (95% CI) | 20.8 (17.6, 23.9) |
| Month 12, mmHg (95% CI) | 15.0 (12.6, 17.4) |
| Absolute change, mmHg (95% CI) | −5.8 (−8.7, −2.8) |
| Relative change, % (95% CI) | −22.5% (−33.6%, −11.4%) |
| p-value | p < 0.001 |
| **Responder analyses at Month 12 (N=32), n (%)** |  |
| **IOP threshold** |  |
| ≤ 21 mmHg | 31 (96.9%) |
| ≤ 18 mmHg | 28 (87.5%) |
| ≤ 15 mmHg | 22 (68.8%) |
| ≥ 20% reduction | 22 (68.8%) |
| ≥ 30% reduction | 13 (40.6%) |
| **Composite responder, IOP threshold without increase in medication** |  |
| ≤ 21 mmHg without increase in medication | 23 (71.9%) |
| ≤ 18 mmHg without increase in medication | 22 (68.8%) |
| ≤ 15 mmHg without increase in medication | 18 (56.2%) |
| ≥ 20% reduction without increase in medication | 16 (50.0%) |
| ≥ 30% reduction without increase in medication | 9 (28.1%) |

CI = confidence interval; IOP = intraocular pressure; SD = standard deviation.
\* Month 1 medication data should be interpreted with caution due to the common clinical practice of temporarily withholding or reducing ocular hypotensive medications postoperatively.
Note: Relative changes are calculated as the mean of per-eye percentage changes.

Mean medication burden as observed was 3.0 (1.3) at baseline (N=47), 2.1 (1.6) at Month 1 (N=42), 2.7 (1.7) at Month 6 (N=37) and 2.8 (1.5) at Month 12 (N=32). Among the 32 eyes evaluable at Month 12, mean medication burden was 2.97 (1.31) at baseline and 2.84 (1.46) at Month 12, a change of −0.12 (−0.50, +0.25), which was not statistically significant (p = 0.50). The proportion of medication-free eyes was 4.3% (2 of 47) at baseline, 10.8% (4 of 37) at Month 6 and 3.1% (1 of 32) at Month 12. The Month 1 proportion (16.7%) was artificially high due to the common clinical practice of temporarily withholding glaucoma medication in the early postoperative period.

Corneal endothelial cell count was available at both baseline and Month 12 in 7 eyes (14.9%), reflecting that real-world specular microscopy is not required by the CREST protocol and not part of routine glaucoma care. In eyes with evaluable ECC data, the mean was 2,002 cells/mm² at baseline and 2,060 cells/mm² at Month 12, a change of +59 cells/mm² (−72, +189; p = 0.31), or +3.9% (−2.4%, +10.1%); individual changes ranged from −7.2% to +9.5%.

### Safety Profile

Eleven of 47 eyes (23.4%) underwent an incisional reoperation during the follow-up period. Eight eyes (17.0%) proceeded to bleb-forming filtration, comprising a gel stent in five, trabeculectomy in two and an ExPRESS shunt in one, at a median of 190 days; three eyes (6.4%) received a glaucoma drainage device, at a median of 350 days. The Kaplan-Meier estimate (95% CI) for freedom from reoperation at 12 months was 78.2% (61.8%, 88.2%) (Table 3, Figure 5). Seventeen eyes remained at risk at 12 months. Twenty-one eyes without a reoperation were censored before 12 months: 17 at a Month 12 visit held between days 277 and 365, 2 at their Month 1 or Month 6 visit, and 2 that had no scheduled follow-up visit. Separately, four eyes (8.5%) underwent cyclophotocoagulation (five procedures) and two eyes (4.3%) underwent laser trabeculoplasty; no eye underwent laser peripheral iridotomy. All seven procedures were performed for inadequate IOP control, in one case with poor adherence to medication noted, and four of them took place within the first 12 months. One of the cyclophotocoagulation procedures preceded an incisional reoperation in the same eye. When any subsequent IOP-lowering procedure was counted as a failure, the Kaplan-Meier estimate of freedom from further intervention at 12 months was 70.1% (52.8%, 82.1%), with 16 eyes at risk at 12 months (Table 3, Figure 6).

**Figure 5.**
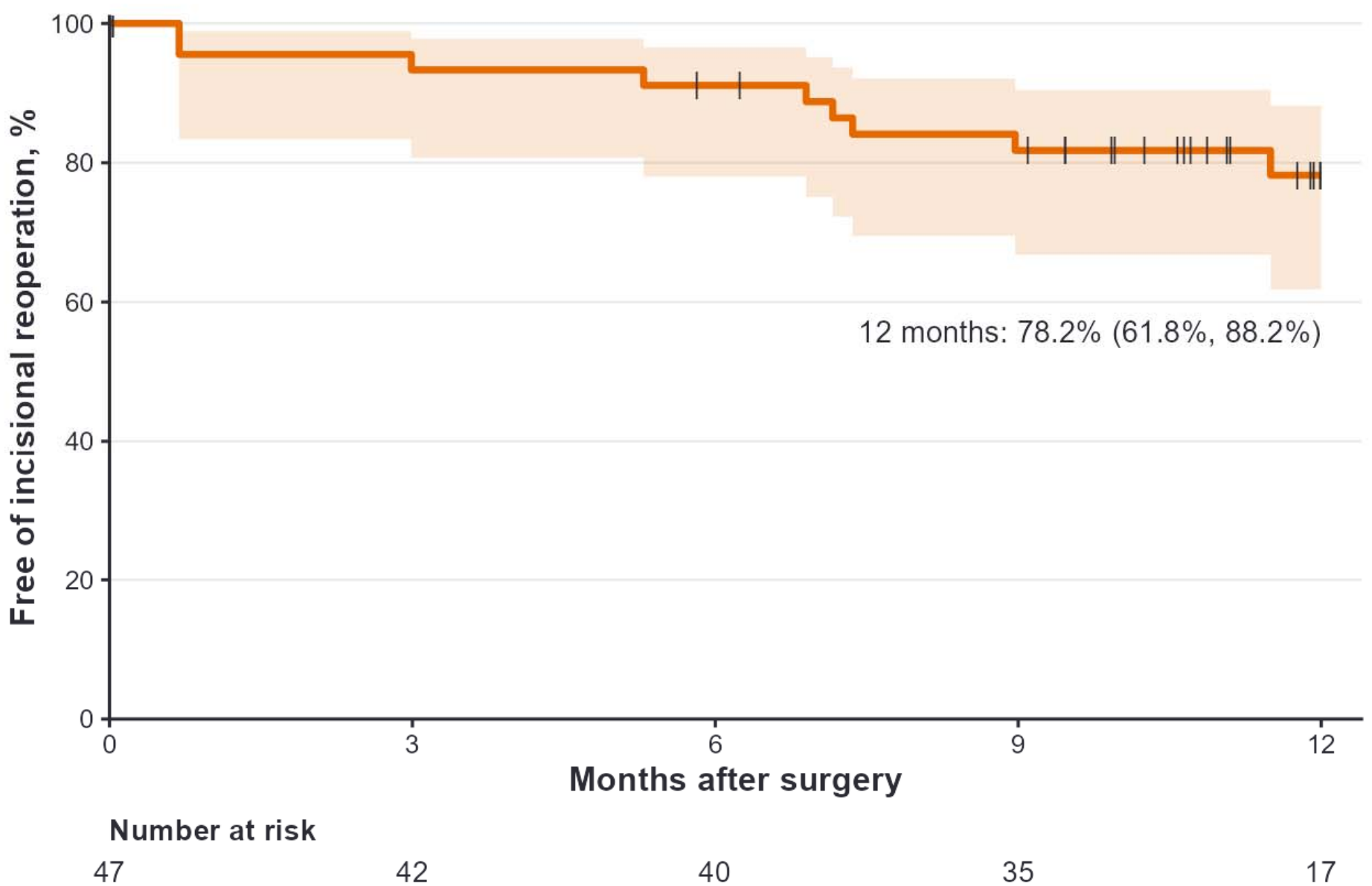
Kaplan-Meier Freedom from Incisional Reoperation. CI = confidence interval; IOP = intraocular pressure. Shaded band shows the 95% CI (complementary log-log). Vertical ticks mark censored eyes. An event is any incisional IOP-lowering procedure (gel stent, trabeculectomy, ExPRESS shunt, or tube shunt). Eyes without an event were censored at their last scheduled study visit; two eyes with no scheduled follow-up visit were censored on day 1. Numbers at risk are shown beneath the time axis.

**Figure 6.**
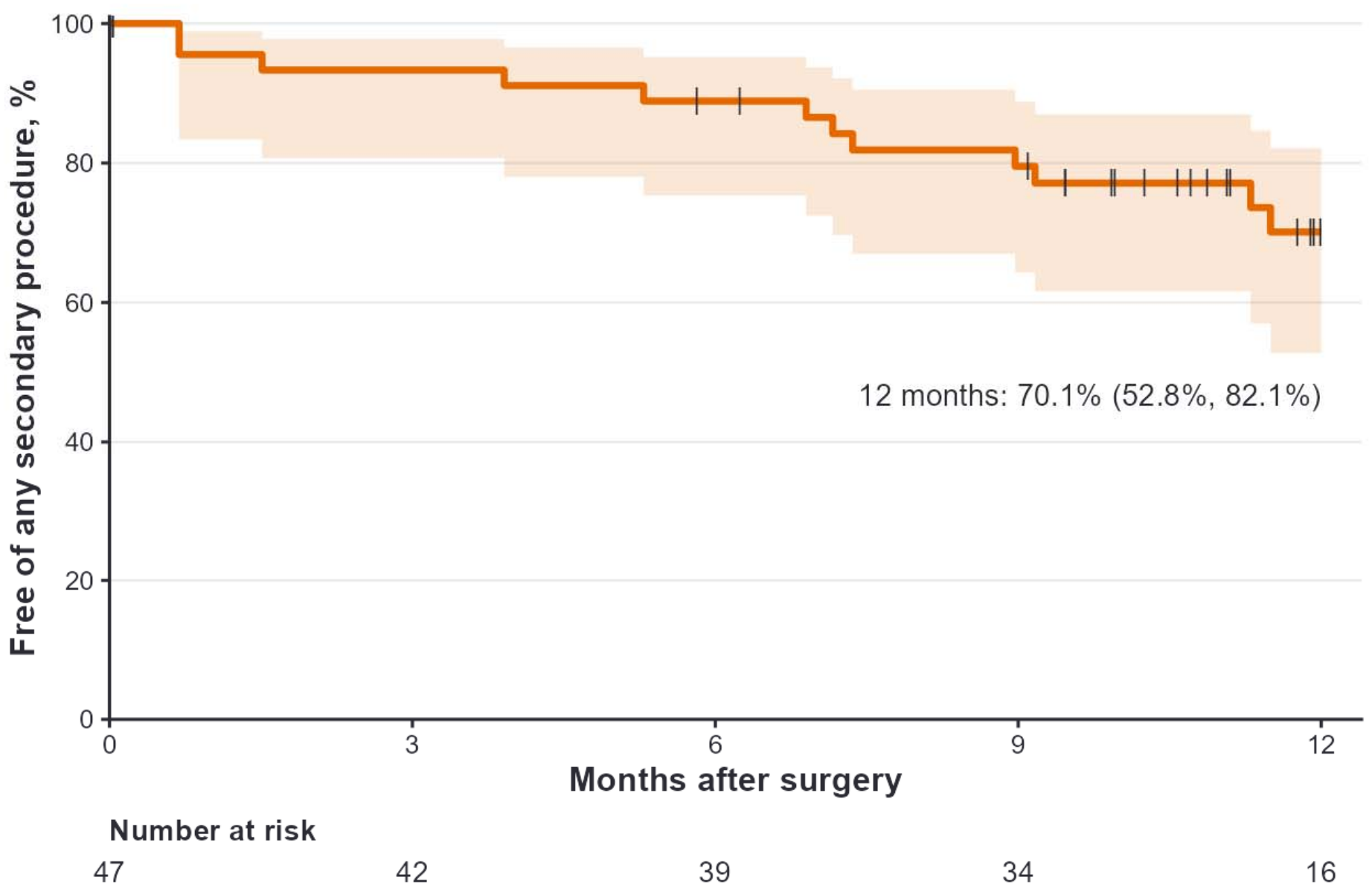
Kaplan-Meier Freedom from Any Secondary IOP-Lowering Procedure. CI = confidence interval; IOP = intraocular pressure. Shaded band shows the 95% CI (complementary log-log). Vertical ticks mark censored eyes. An event is any subsequent IOP-lowering procedure: incisional surgery (Figure 5), in addition to cyclophotocoagulation, laser trabeculoplasty, or laser peripheral iridotomy. Anterior chamber paracentesis for acute IOP elevation was not counted. Eyes without an event were censored at their last scheduled study visit (Month 1, 6 or 12); two eyes with no scheduled follow-up visit were censored on day 1. Numbers at risk are shown beneath the time axis.

**Table 3.**
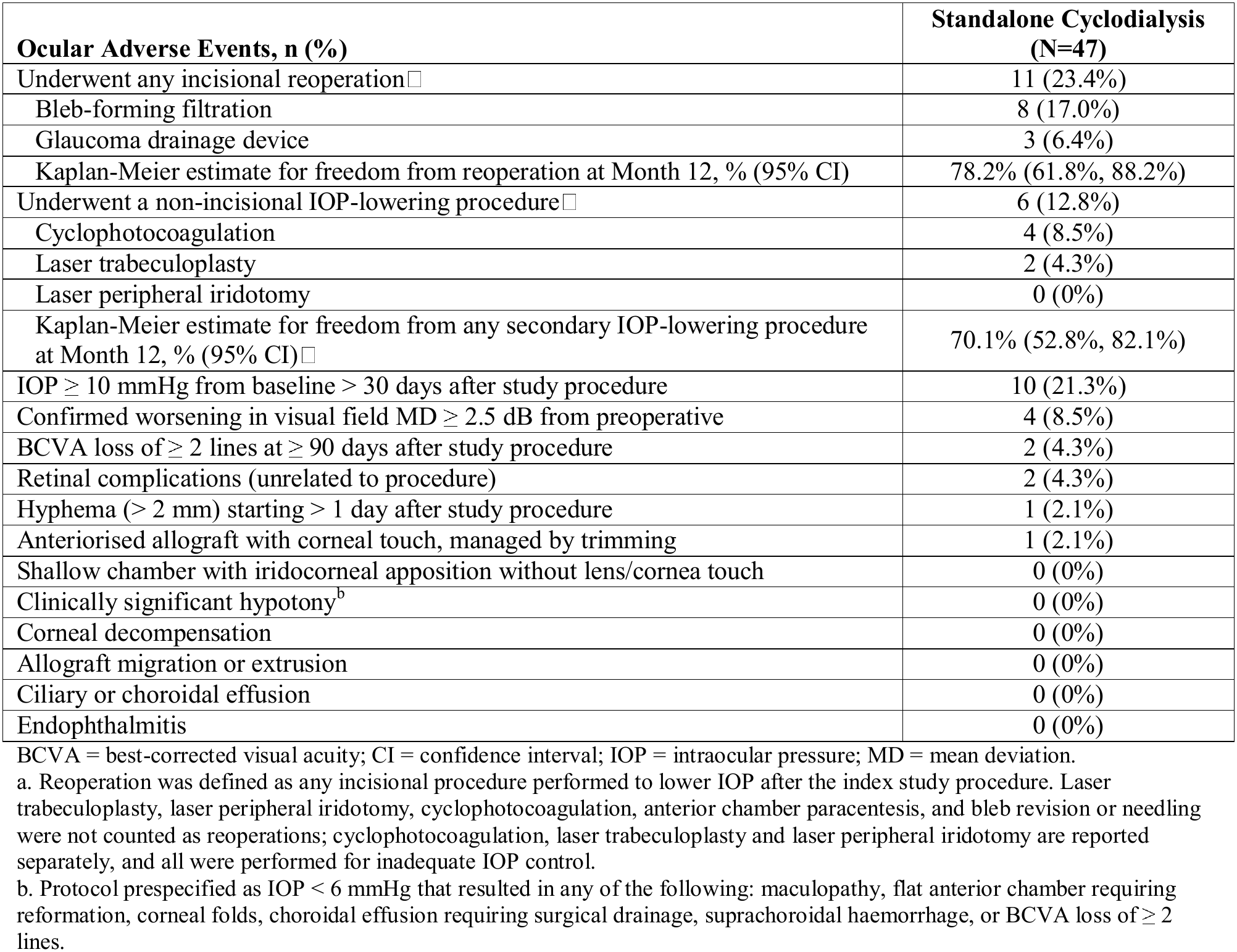
Summary of Ocular Adverse Events.

| Ocular Adverse Events, n (%) | Standalone Cyclodialysis (N=47) |
|---|---|
| Underwent any incisional reoperation□ | 11 (23.4%) |
| Bleb-forming filtration | 8 (17.0%) |
| Glaucoma drainage device | 3 (6.4%) |
| Kaplan-Meier estimate for freedom from reoperation at Month 12, % (95% CI) | 78.2% (61.8%, 88.2%) |
| Underwent a non-incisional IOP-lowering procedure□ | 6 (12.8%) |
| Cyclophotocoagulation | 4 (8.5%) |
| Laser trabeculoplasty | 2 (4.3%) |
| Laser peripheral iridotomy | 0 (0%) |
| Kaplan-Meier estimate for freedom from any secondary IOP-lowering procedure at Month 12, % (95% CI)□ | 70.1% (52.8%, 82.1%) |
| IOP ≥ 10 mmHg from baseline > 30 days after study procedure | 10 (21.3%) |
| Confirmed worsening in visual field MD ≥ 2.5 dB from preoperative | 4 (8.5%) |
| BCVA loss of ≥ 2 lines at ≥ 90 days after study procedure | 2 (4.3%) |
| Retinal complications (unrelated to procedure) | 2 (4.3%) |
| Hyphema (> 2 mm) starting > 1 day after study procedure | 1 (2.1%) |
| Anteriorised allograft with corneal touch, managed by trimming | 1 (2.1%) |
| Shallow chamber with iridocorneal apposition without lens/cornea touch | 0 (0%) |
| Clinically significant hypotony<sup>b</sup> | 0 (0%) |
| Corneal decompensation | 0 (0%) |
| Allograft migration or extrusion | 0 (0%) |
| Ciliary or choroidal effusion | 0 (0%) |
| Endophthalmitis | 0 (0%) |

BCVA = best-corrected visual acuity; CI = confidence interval; IOP = intraocular pressure; MD = mean deviation.
a. Reoperation was defined as any incisional procedure performed to lower IOP after the index study procedure. Laser trabeculoplasty, laser peripheral iridotomy, cyclophotocoagulation, anterior chamber paracentesis, and bleb revision or needling were not counted as reoperations; cyclophotocoagulation, laser trabeculoplasty and laser peripheral iridotomy are reported separately, and all were performed for inadequate IOP control.
b. Protocol prespecified as IOP < 6 mmHg that resulted in any of the following: maculopathy, flat anterior chamber requiring reformation, corneal folds, choroidal effusion requiring surgical drainage, suprachoroidal haemorrhage, or BCVA loss of ≥ 2 lines.

There were no cases of clinically significant hypotony (IOP < 6 mmHg with clinical complications) at any time during follow-up, and no cases of ciliary or choroidal effusion, allograft extrusion, corneal endothelial decompensation or endophthalmitis.

Protocol-specified IOP elevations ≥ 10 mmHg from baseline occurring > 30 days postoperatively were reported in 10 eyes (21.3%), predominantly managed medically with adjustment of IOP-lowering medication. Hyphaema > 2 mm persisting beyond the first postoperative day occurred in 1 eye (2.1%), in a patient taking a systemic anticoagulant, and resolved with conservative management without medication or surgery. Incidental retinal complications over the 12-month follow-up period occurred in 2 eyes (4.3%): 1 retinal tear and 1 retinal detachment. Both were assessed by the investigator as unrelated to the allograft or study procedure. No eye in this cohort had a shallow chamber with iridocorneal apposition.

Postoperative gonioscopy was performed at least once in 30 eyes (63.8%). Among those eyes, the allograft was recorded as optimally positioned in 27 (90.0%), and the cleft was open around the allograft in 3 (10.0%). Partial synechiae around or over the allograft were recorded in 2 (6.7%) eyes. None had total synechiae. Findings are not mutually exclusive.

One eye had an adverse event of corneal endothelial touch by the allograft. This eye had previously undergone laser trabeculoplasty and three angle-based procedures and had subsequently received cyclophotocoagulation and a gel stent; the finding was attributed by the investigator to post-operative changes following those reinterventions. Surgical trimming of the allograft was performed, and the event was reported as resolved without sequelae. No eye had an adverse event coded as allograft migration or extrusion.

No eye experienced any of the following protocol-prespecified AEs: chronic ocular pain, flat anterior chamber requiring reformation, macular oedema, hypotonic maculopathy, choroidal folds, other maculopathy, vitreous haemorrhage, wound dehiscence, or chronic anterior uveitis.

## 4. Discussion

In this CREST subgroup of 47 eyes uncontrolled after prior trabecular intervention, standalone allograft-reinforced cyclodialysis of the complementary uveoscleral pathway was associated with a 25.0% IOP reduction at 12 months (p < 0.001), achieved without an increase in medication burden, in an advanced population (61.7% on three or more medications, mean visual field mean deviation −11.7 dB, 95.7% pseudophakic). Kaplan-Meier freedom from reoperation was 78.2%, or 70.1% when any further IOP-lowering procedure was counted as a failure, and there were no cases of clinically significant hypotony, which was the failure mode of unreinforced cyclodialysis.

### The Physiological Rationale

The rationale for this approach is physiological. Conventional and uveoscleral outflow are distinct systems with distinct rate-limiting resistances^2,3^. Trabecular procedures are constrained by resistance distal to the trabecular meshwork and by episcleral venous pressure, which together impose a practical IOP floor in the mid-teens and cannot be overcome by re-operating on the same route^7,8^. In randomised studies, trabecular procedures have been shown to add only about 1.6 to 2.3 mmHg over cataract surgery, and roughly three-quarters of the reduction seen in combined procedures comes from the lens extraction^9^. In a pseudophakic eye that has already failed a trabecular intervention, both the lens contribution and much of the trabecular capacity have already been used. Redirecting the surgical effort to the untouched, comparatively pressure-independent uveoscleral pathway is therefore a mechanistically distinct escalation rather than a repeat of a route that has already failed^4,14^. The preserved effect in the stricter non-laser subgroup (22.5% reduction, paired N=25) supports the interpretation that benefit derives from engaging a new pathway rather than from regression of prior laser effect.

Because it is ab-interno and leaves the conjunctiva undisturbed, allograft-reinforced cyclodialysis preserves the option of future bleb-forming surgery, whose success is degraded by prior conjunctival manipulation and scarring^1,10^. In a moderate-to-advanced cohort, many of whom will eventually need filtration, deferring conjunctival disturbance while still lowering IOP and avoiding added drops is a practical advantage in sequencing.

### Contextual Comparison with Trabecular MIGS

Cross-study comparisons with published trabecular MIGS series should be interpreted with substantial caution, because baseline characteristics such as prior surgery, baseline IOP, medication burden and lens status differ materially across cohorts and no head-to-head data exist. IRIS Registry analyses have reported cumulative reoperation rates of 15%–24% at 2 years after standalone MIGS procedures^20^. In the present cohort, 11 of 47 eyes (23.4%) underwent reoperation during the observation window. For context, a prospective single-arm study of standalone trabecular micro-bypass in eyes uncontrolled by prior surgical or medical therapy reported a mean diurnal IOP reduction of 5.9 mmHg at 12 months from a baseline of 23.4 mmHg on a mean of 3.1 medications, and 5.5 mmHg in the subgroup with prior failed surgery^21^. In the present cohort, the 32 eyes contributing a paired Month 12 measurement had a baseline IOP of 20.8 mmHg on a mean of 3.0 medications and a mean reduction of 6.1 mmHg. Differences in IOP ascertainment, baseline pressure and study design preclude direct comparison. These figures are offered as contextual background only, and direct comparative trials would be required to establish the relative position of allograft-reinforced cyclodialysis against trabecular MIGS.

### Positioning in the Surgical Algorithm

Allograft-reinforced cyclodialysis produced meaningful standalone IOP reduction in eyes with prior trabecular intervention while leaving the conjunctiva intact for future filtering surgery. In the surgical sequence it fits well between a second trabecular attempt and bleb-forming surgery.

## Limitations

First, this is a single-arm, non-randomised sub-study of a real-world evidence study of consecutive, prospectively followed subjects, in which each patient was treated with the postoperative standard of care as applied by their treating physician. Selection bias and confounding are inherent to this observational design and cannot be fully controlled. Second, the efficacy analysis is confined to the 32 eyes that contributed a Month 12 measurement before any subsequent IOP-lowering procedure, and the definition of disease control is not protocolised but assessed individually by the treating physicians’ discretion. Third, no medication washout was performed, so IOP and medication outcomes are interdependent. The composite responder endpoint (IOP threshold met without added medication) is the most conservative reading and is reported for this reason. Fourth, the “prior trabecular intervention” population was heterogeneous, dominated by laser trabeculoplasty. The non-laser sensitivity analysis mitigates this but cannot eliminate it. Fifth, the Kaplan-Meier estimates confidence intervals are relatively wide; the durability of overall success therefore could benefit from longer follow-up. Finally, corneal endothelial data are sparse and available in only a fraction of patients, as specular microscopy is not standard of care in interventional glaucoma patients.

## Data Availability

The data that support the findings of this study are available from the corresponding author upon reasonable request.

## Acknowledgements

This study is registered in the ClinicalTrials.gov trial registry (identifier: NCT05506423; date of registration: 18 August 2022).

## Author Contributions

Conceptualisation: Tsontcho Ianchulev. Methodology and formal analysis: Tsontcho Ianchulev. Investigation and data acquisition: Gus Gazzard, Hari Jayaram, Reena Garg, Robert Stamper, James Tsai and I. Paul Singh. Writing, original draft preparation: Tsontcho Ianchulev. Writing, review and editing: all authors. Supervision: Tsontcho Ianchulev. All authors read and approved the final manuscript.

